# A preliminary study on serological assay for severe acute respiratory syndrome coronavirus 2 (SARS-CoV-2) in 238 admitted hospital patients

**DOI:** 10.1101/2020.03.06.20031856

**Authors:** Lei Liu, Wanbing Liu, Yaqiong Zheng, Xiaojing Jiang, Guomei Kou, Jinya Ding, Qiongshu Wang, Qianchuan Huang, Yinjuan Ding, Wenxu Ni, Wanlei Wu, Shi Tang, Li Tan, Zhenhong Hu, Weitian Xu, Yong Zhang, Bo Zhang, Zhongzhi Tang, Xinhua Zhang, Honghua Li, Zhiguo Rao, Hui Jiang, Xingfeng Ren, Shengdian Wang, Shangen Zheng

## Abstract

**Background:** The outbreak of the recently emerged novel corona virus disease 2019 (COVID-19) poses a challenge for public health laboratories. We aimed to evaluate the diagnostic value of serological assay for SARS-CoV-2.

**Methods:** A newly-developed ELISA assay for IgM and IgG antibodies against N protein of SARS-CoV-2 were used to screen the serums of 238 admitted hospital patients with confirmed or suspected SARS-CoV-2 infection from February 6 to February 14, 2020. SARS-CoV-2 RNA was detected by real time RT-PCR on pharyngeal swab specimens.

**Findings:** Of the 238 patients, 194 (81.5%) were detected to be antibody (IgM and/or IgG) positive, which was significantly higher than the positive rate of viral RNA (64.3%). There was no difference in the positive rate of antibody between the confirmed patients (83.0%, 127/153) and the suspected patients (78.8%, 67/85) whose nucleic acid tests were negative. After the patients were defined to the different stages of disease based on the day when the test samples were collected, the analysis results showed that the antibody positive rates were very low in the first five days after initial onset of symptoms, and then rapidly increased as the disease progressed. After 10 days, the antibody positive rates jumped to above 80% from less than 50%. On the contrary, the positive rates of viral RNA kept above 60% in the first 11 days after initial onset of symptoms, and then rapidly decreased. In addition, half of the suspected patients with symptoms for 6-10 days were detected to be antibody positive.

**Interpretation:** The suspected patients were most likely infected by SARS-CoV-2. Before the 11th day after initial onset of symptoms, nucleic acid test is important for confirmation of viral infection. The combination of serological assay can greatly improve the diagnostic efficacy. After that, the diagnosis for viral infection should be majorly dependent on serological assay.

## Introduction

A novel betacoronavirus ^1^ named severe acute respiratory syndrome coronavirus 2 (SARS-CoV-2) causes a recent cluster cases of respiratory illness named corona virus disease 2019 (COVID-19) in multiple regions of the world, and it leads to a serious public health problem especially in Wuhan city, Hubei province, China since December 2019. ^2^ By March 1, 2020, more than 80,000 confirmed cases have been identified globally both in China and other 58 countries spanning Asia, Europe, Oceania, North and South America, and Northeast Africa. It is evidenced that SARS-CoV-2 can transmit rapidly from person to person, which is evidently found in hospital and family settings. ^3-5^

The SARS-CoV-2 is the seventh member of enveloped RNA coronaviruses (CoVs). ^6-8^ Sequence and phylogenetic tree of CoVs analysis indicates that SARS-CoV-2 is genetically distinct from SARS-CoV and is more closely related to bat-SL-CoV ZC45 and bat-SL-CoV ZXC21. ^1^ SARS-CoV-2 owns a similar receptor-binding domain structure to that of SARS-CoV. ^1^ A typical CoV contains four main structural proteins: spike (S), membrane (M), envelope (E), and nucleocapsid (N) proteins. The S protein homotrimers are required for attachment to host receptors, ^9^and both the M protein and the E protein play important roles in virus assembly. ^10,11^ The N protein is responsible for packaging the encapsidated genome into virions, ^12,13^ and acts as a viral RNA silencing suppressor that is beneficial for the viral replication. ^14^ Furthermore, the N protein has high immunogenic activity and is profusely overexpressed during infection, ^15^ indicating that N protein should be a potential source of a diagnostic antigen for detecting SARS-CoV-2 infection. Many diagnostic methods based on the N protein have been developed for SARS-CoV detection. ^16-18^ In addition, different CoVs possess special structural and accessory proteins, such as HE protein, 3a/b protein, and 4a/b protein. ^19^

Both nucleic acid test and serological assay are commonly used for infectious disease screening and diagnosis. In the present case of SARS-CoV-2, nucleic acid test has been being routinely used to detect causative viruses from respiratory secretions by real-time RT-PCR in China. However, the nucleic acid tests appeared to have a high false negative rate because of several unavoidable reasons, including the sensitivity of the detection kits that have not been well assessed, sampling location and technique, etc. A large number of clinically-suspected patients, whose nucleic acid tests were negative, are unable to get timely confirmed-diagnosis and hospital treatment, which potentially promotes the spread of SARS-CoV-2 and leads to a rapid disease progress of the suspected patients.

In this study, a newly-developed IgM and IgG antibody detecting Enzyme-linked immunosorbent assays (ELISA) based on a recombinant fragment of SARS-CoV-2 N protein were used to detect IgM and IgG against SARS-CoV-2 in serum of 238 admitted hospital patients with confirmed or suspected SARS-CoV-2 infection. The results strongly indicated that the suspected patients were infected. We also analyze the diagnostic value of the IgM and IgG testing in COVID-19, even in the early stage of disease.

## Methods

### Patients and samples

All consecutive patients (n=238) with confirmed or suspected SARS-CoV-2 infection who have been tested by real-time RT-PCR for viral infection and were being treated in General Hospital of Central Theater Command of PLA from February 6 to February 14, 2020, were enrolled. The general information (age, sex, vital signs, coexisting disorders), clinical, laboratory, and radiological characteristics data of the patients on admission were extracted from electronic medical records. Among the 238 recruited patients, 153 patients were laboratory-confirmed cases, who were tested positive for viral RNA by real time RT-PCR assay on pharyngeal swab specimens, and the remaining 85 patients having negative results for real time RT-PCR assay were clinically diagnosed as highly-suspected cases according to the notice on the issuance of strategic guidelines for diagnosis and treatment of COVID-19. ^20^ The serum samples were collected once from each recruited patient. Meanwhile, the serum samples from 70 ordinary patients and 50 healthy blood donors were randomly selected as the controls. The study was approved by the Hospital Ethics Committee and written informed consent was waived for emerging infectious diseases.

### Real time Reverse Transcription Polymerase Chain Reaction (RT-PCR) Assay

Pharyngeal swab specimens were collected from patients and placed into a collection tube with 200 μL of virus preservation solution. Total RNA was extracted using the respiratory sample RNA isolation kit (Shuoshi, Shanghai, China). After vortex, 50 μL of cell lysates were transferred into another collection tube. The collection tube was centrifugated at 1000 rpm/min for 5 min after standing at room temperature for 10 minutes. 5 μL RNA was prepared and used for real time RT-PCR.

Real time RT-PCR was performed using the nucleic acid testing kit (Daan, Guangzhou, China) for SARS-CoV-2 detection. The open reading frame 1ab (*ORF1ab*) and nucleocapsid protein (N) were simultaneously selected as the two target genes. The human *GAPDH* gene was used as an internal control. The specific primers and probes set for *ORF1ab* and N were as follows: *ORF1ab-*forward primer 5’-ACCTTCTCTTGCCACTGTAGC-3’; *ORF1ab-*reverse primer 5’-AGTATCAACCATATCCAACCATGTC-3’; and the probe 5’-FAM-ACGCATCACCCAACTAGCAGGCATAT-BHQ1-3’; N*-*forward primer 5’-TTCAAGAAATTCAACTCCAG-3’; N*-*reverse primer 5’-AGCAGCAAAGCAAGAGCAGCATC-3’; and the probe 5’-VIC-TCCTGCTAGAATGGCTGGCAATGGCG-BHQ1-3’. The real time RT-PCR experiment was thoroughly performed according to kit’s instructions. The reaction mixture contains 17 μL of reaction buffer A, 3 μL of reaction buffer B, and 5 μL RNA template. The real time RT-PCR assay was performed under the following conditions: incubation at 50 °C for 15 min and 95 °C for 15 min, 45 cycles of denaturation at 94 °C for 15 s, and extending and collecting fluorescence signal at 55 °C for 45 s. A cycle threshold value (Ct-value) ≤ 40 was defined as a positive test result, and a Ct-value > 40 was defined as a negative test.

### Enzyme-Linked Immunosorbent Assay (ELISA)

Serological assay was performed using Enzyme-Linked Immunosorbent Assays kit (Lizhu, Zhuhai, China), which was established for detecting IgM or IgG antibody against N protein of SARS-CoV-2. For IgM detection, ELISA plates were previously coated with mouse anti-human IgM (μ chain) monoclonal antibody. 100 μL diluted (1:100) serum sample was added into the pre-coated plates with three replicating wells for each sample and incubated at 37 °C for 1 h. The heat-inactivated positive and negative serum were included on each plate. After washing, 100 μL horse radish peroxidase (HRP) conjugated recombinant (rN) protein of SARS-CoV-2 was added. Then the plate was incubated at 37 °C for 30 min followed by washing. 50 μL of TMB substrate solution and 50 μL of the corresponding buffer were added and incubated at 37 °C for 15 min. The reaction was terminated by adding 50 μL of 2 M sulfuric acid, and the absorbance value at 450 nm (A_450_) was determined. The cut off value was calculated by sum of 0.100 and average A_450_ of negative control replicates. A_450_ less than cut off value was defined as a negative test, and A_450_ greater than or equal to cut off value was defined as a positive test.

For IgG detection, ELISA plates were previously coated with rN protein. 5 μL serum sample diluted with 100 μL dilution buffer were added into the plates. After incubation and washing, HRP-conjugated mouse anti-human IgG monoclonal antibody was added into the plates for detection. The other operation steps were performed as described in the above IgM detection. The cut off value was calculated by the sum of 0.130 and average A_450_ of negative control replicates. A_450_ less than cut off value was defined as a negative test, and A_450_ greater than or equal to cut off value was defined as a positive test.

### Statistical analysis

Continuous variables were described as the means and standard deviations or medians and interquartile ranges (IQR) values. Categorical variables were expressed as the counts and percentages. Independent group t tests were applied to continuous variables that were normally distributed; otherwise, the Mann-Whitney test was used. Categorical variables were compared using the chi-square tests, while the Fisher exact test was used when the data were limited. Statistical analyses were performed using Statistical Package for the Social Sciences (SPSS) version 22.0 software. A two-sided α of less than 0.05 was considered statistically significant.

## Results

### Demographics and patient characteristics

The serum samples were collected from 238 admitted hospital patients with confirmed or suspected SARS-CoV-2 infection in General Hospital of Central Theater Command of PLA from February 6 to February 14, 2020. The clinical characteristics of the patients were shown in Table 1. The median age was 55 years (IQR, 38.3-65), and 138 (58.0%) of the patients were men. Hypertension (63 [26.5%]), diabetes (25 [10.5%]), and cardiovascular disease (24 [10.1%]) were the most common coexisting disorders. Of these patients, the most common symptoms at illness onset were fever (206 [86.6%]), dry cough (128 [53.8%]), and fatigue (78 [32.8%]). A small number of patients possessed the symptoms of abdominal pain (1 [0.4%]), vomiting (3 [1.3%]), and dizziness (4 [1.7%]). On admission, leucocytes were below the normal range in 41 (17.2%) patients and above the normal range in 21 (8.8%) patients. 125 (52.5%) patients had lymphocytes below the normal range and no patients were found to have lymphocytes above the normal range. Neutrophils were below the normal range in 28 (11.8%) patients and above the normal range in 32 (13.4%) patients. According to CT, 235 (98.7%) patients showed ground-glass opacity and/or patchy shadowing.

**Table 1.**
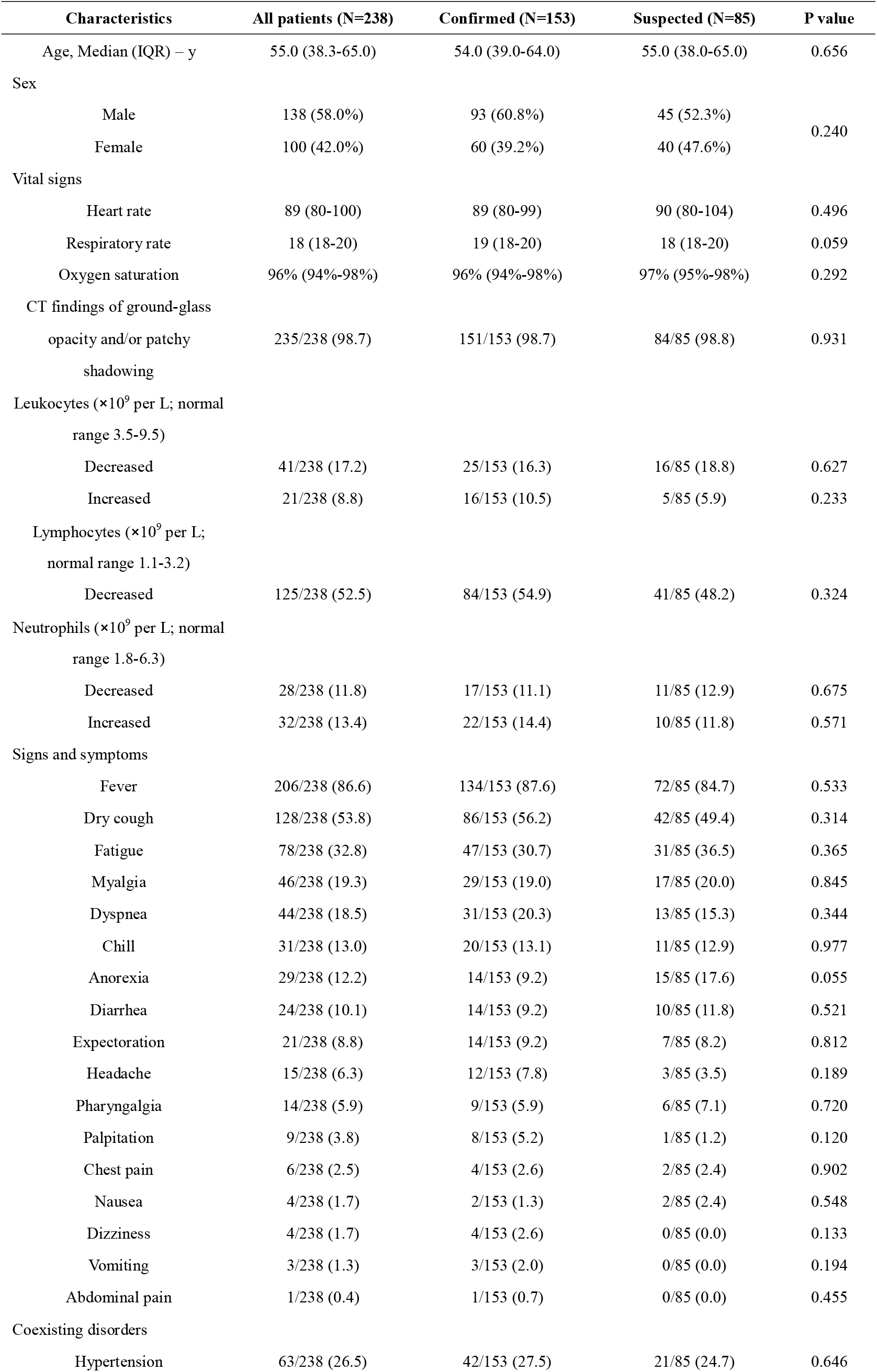
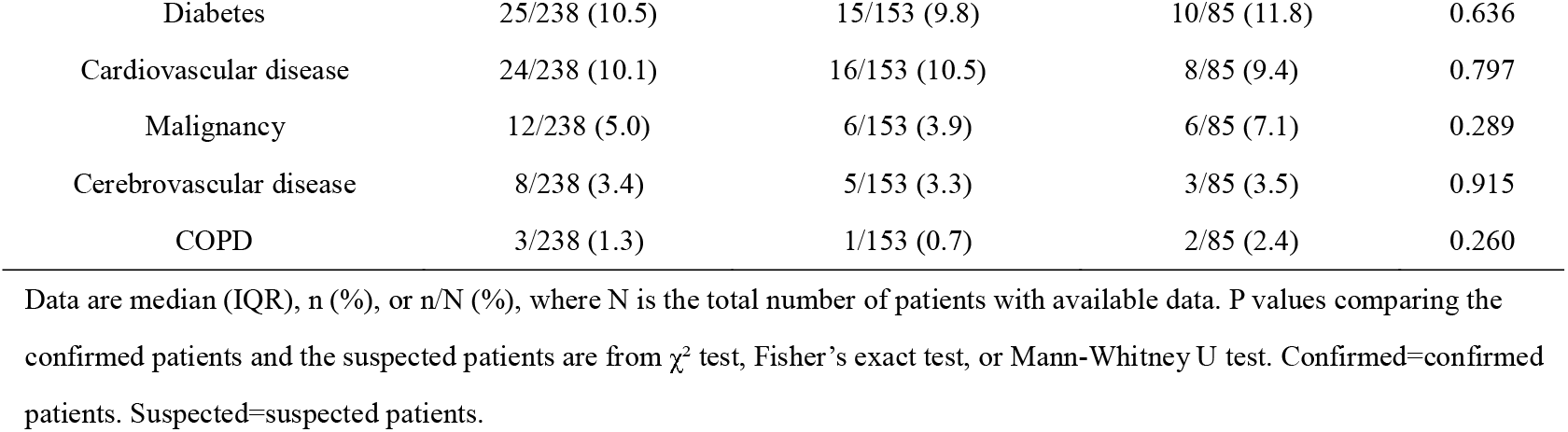
Demographic and baseline characteristics of 238 enrolled patients

| Characteristics | All patients (N=238) | Confirmed (N=153) | Suspected (N=85) | P value |
| --- | --- | --- | --- | --- |
| Age, Median (IQR) – y | 55.0 (38.3-65.0) | 54.0 (39.0-64.0) | 55.0 (38.0-65.0) | 0.656 |
| Sex |  |  |  |  |
| Male | 138 (58.0%) | 93 (60.8%) | 45 (52.3%) | 0.240 |
| Female | 100 (42.0%) | 60 (39.2%) | 40 (47.6%) |  |
| Vital signs |  |  |  |  |
| Heart rate | 89 (80-100) | 89 (80-99) | 90 (80-104) | 0.496 |
| Respiratory rate | 18 (18-20) | 19 (18-20) | 18 (18-20) | 0.059 |
| Oxygen saturation | 96% (94%-98%) | 96% (94%-98%) | 97% (95%-98%) | 0.292 |
| CT findings of ground-glass opacity and/or patchy shadowing | 235/238 (98.7) | 151/153 (98.7) | 84/85 (98.8) | 0.931 |
| Leukocytes ($\times 10^9$ per L; normal range 3.5-9.5) | | | | |
| Decreased | 41/238 (17.2) | 25/153 (16.3) | 16/85 (18.8) | 0.627 |
| Increased | 21/238 (8.8) | 16/153 (10.5) | 5/85 (5.9) | 0.233 |
| Lymphocytes ($\times 10^9$ per L; normal range 1.1-3.2) | | | | |
| Decreased | 125/238 (52.5) | 84/153 (54.9) | 41/85 (48.2) | 0.324 |
| Neutrophils ($\times 10^9$ per L; normal range 1.8-6.3) | | | | |
| Decreased | 28/238 (11.8) | 17/153 (11.1) | 11/85 (12.9) | 0.675 |
| Increased | 32/238 (13.4) | 22/153 (14.4) | 10/85 (11.8) | 0.571 |
| Signs and symptoms |  |  |  |  |
| Fever | 206/238 (86.6) | 134/153 (87.6) | 72/85 (84.7) | 0.533 |
| Dry cough | 128/238 (53.8) | 86/153 (56.2) | 42/85 (49.4) | 0.314 |
| Fatigue | 78/238 (32.8) | 47/153 (30.7) | 31/85 (36.5) | 0.365 |
| Myalgia | 46/238 (19.3) | 29/153 (19.0) | 17/85 (20.0) | 0.845 |
| Dyspnea | 44/238 (18.5) | 31/153 (20.3) | 13/85 (15.3) | 0.344 |
| Chill | 31/238 (13.0) | 20/153 (13.1) | 11/85 (12.9) | 0.977 |
| Anorexia | 29/238 (12.2) | 14/153 (9.2) | 15/85 (17.6) | 0.055 |
| Diarrhea | 24/238 (10.1) | 14/153 (9.2) | 10/85 (11.8) | 0.521 |
| Expectoration | 21/238 (8.8) | 14/153 (9.2) | 7/85 (8.2) | 0.812 |
| Headache | 15/238 (6.3) | 12/153 (7.8) | 3/85 (3.5) | 0.189 |
| Pharyngalgia | 14/238 (5.9) | 9/153 (5.9) | 6/85 (7.1) | 0.720 |
| Palpitation | 9/238 (3.8) | 8/153 (5.2) | 1/85 (1.2) | 0.120 |
| Chest pain | 6/238 (2.5) | 4/153 (2.6) | 2/85 (2.4) | 0.902 |
| Nausea | 4/238 (1.7) | 2/153 (1.3) | 2/85 (2.4) | 0.548 |
| Dizziness | 4/238 (1.7) | 4/153 (2.6) | 0/85 (0.0) | 0.133 |
| Vomiting | 3/238 (1.3) | 3/153 (2.0) | 0/85 (0.0) | 0.194 |
| Abdominal pain | 1/238 (0.4) | 1/153 (0.7) | 0/85 (0.0) | 0.455 |
| Coexisting disorders |  |  |  |  |
| Hypertension | 63/238 (26.5) | 42/153 (27.5) | 21/85 (24.7) | 0.646 |
| Diabetes | 25/238 (10.5) | 15/153 (9.8) | 10/85 (11.8) | 0.636 |
| Cardiovascular disease | 24/238 (10.1) | 16/153 (10.5) | 8/85 (9.4) | 0.797 |
| Malignancy | 12/238 (5.0) | 6/153 (3.9) | 6/85 (7.1) | 0.289 |
| Cerebrovascular disease | 8/238 (3.4) | 5/153 (3.3) | 3/85 (3.5) | 0.915 |
| COPD | 3/238 (1.3) | 1/153 (0.7) | 2/85 (2.4) | 0.260 |

Data are median (IQR), n (%), or n/N (%), where N is the total number of patients with available data. P values comparing the confirmed patients and the suspected patients are from $\chi^2$ test, Fisher's exact test, or Mann-Whitney U test. Confirmed=confirmed patients. Suspected=suspected patients.

According to the positive or negative results of real time RT-PCR assay for pharyngeal swab specimens, the enrolled patients were divided into two groups: the confirmed group and the suspected group. There were no statistical differences of baseline characteristics of the two groups patients.

### Performance and validation of ELISA assays for viral specific IgM and IgG antibodies

Each serum sample of 238 patients were respectively tested for IgM and IgG antibodies against SARS-CoV-2 by using newly-developed ELISA kits based on N protein of SARS-CoV-2. The IgM and/or IgG could be detected in 194 serum samples, and the positive rate (81.5%) was significantly higher than that of SARS-CoV-2 RNA detected by real time RT-PCR, which was 64.3% (153/238) (Fig.1A). Importantly, there were no difference in positive rates of IgM and/or IgG between the confirmed patients (83.0%, 127/153) and the suspected patients (78.8%, 67/85) (Fig.1B), suggesting that the clinically suspected patients, who were viral RNA negative detected by RT-PCR, were mostly infected.

**Figure 1.**
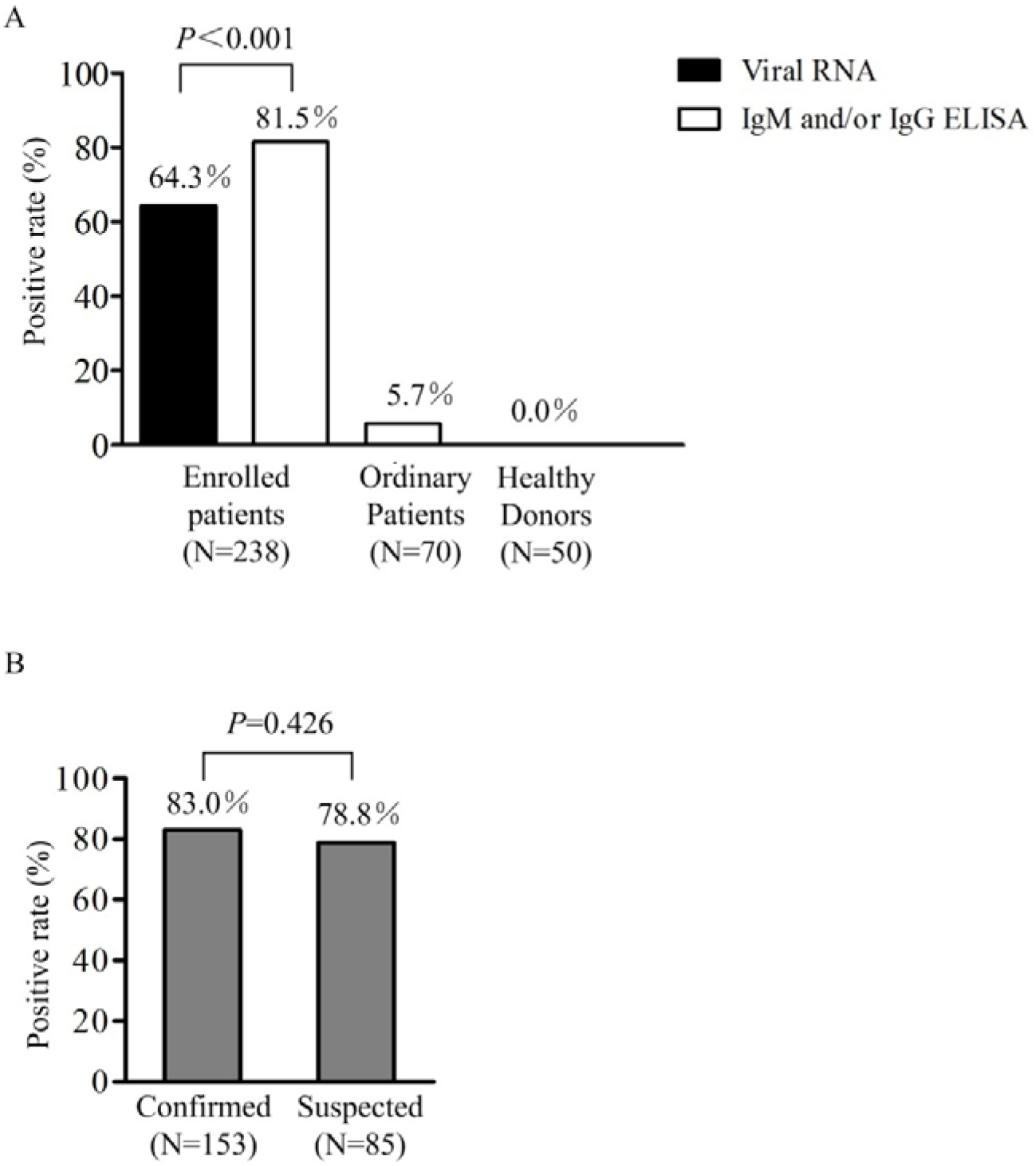
Positive rate of viral RNA and antibody in different samples. A) The positive rate of viral RNA (black column) and antibody (white column) in 238 enrolled patients (two columns on the left), as well as the positive rate of antibody in ordinary patients and healthy donors (two columns on the right). B) Comparison of positive rate of antibody between the laboratory-confirmed (left) and highly-suspected patients (right). Results were compared by chi-square tests.

To verify the specificity of the ELISA assays, the serum samples from 70 randomly-selected ordinary patients and 50 healthy blood donors were simultaneousl detected. Four samples from the ordinary patients were identified as antibody positive (including one dual positive sample, two IgM-positive samples, and one IgG-positive sample) and no positive was found in the samples of healthy blood donors (Fig.1A). These results confirmed the specificity of the IgG and IgM ELISA assays.

### Dynamic analysis of ELISA and RT-PCR assays

To study the diagnostic value of ELISA assay for virus-specific antibodies, especially in the early stage of the disease, we tried to analyze the positive rates of ELISA and RT-PCR assays in the different stages of disease. To this end, each patient was assigned to different days after initial onset of symptoms based on the time when the pharyngeal swab specimen was detected to be positive or the last recoded detection was still negative, or the blood was collected. The positive rates of viral RNA, IgM and/or IgG were compared in every day after initial onset of symptoms (Table 2). Due to a small number of samples in each individual day, we pooled the samples in which the positive rates were similar in consecutive days. Thus, the disease process were divided into five phases of 0-5, 6-10,11-12, 13-15 days and more than 16 days after initial onset of symptoms (Table 3 & Fig.2). The data showed that positive rates of IgM and/or IgG were very low in the first five days after initial onset of symptoms because there was no antibody produced in most of patients in this early stage, and then rapidly increased as the disease progressed. Day 11 after initial onset of symptoms is a key time point because the positive rates of IgM and/or IgG jumped to above 80% from less than 50% at this time point. The dynamic pattern is consistent with SARS-CoV infection. ^21^ On the contrary, the real-time RT-PCR was more effective for detecting SARS-CoV-2 infection than ELISA in the early stage of disease. The positive rate of viral RNA detected by RT-PCR was maintained above 60% in the first 11 days after initial onset of symptoms, and then rapidly decreased with the rapid increase of positive rate of antibodies. These results demonstrated that ELISA-based IgM and/or IgG detection should be used as a major viral diagnostic test for the patients with symptoms for more than 10 days. Because about 50% clinically-suspected patients with symptoms for 6-10 days were detected to be positive by ELISA-based IgM and/or IgG detection (Table 4), the combination of ELISA and RT-PCR assays will greatly improve the detection efficacy, even in the early stage of COVID-19 infection.

**Figure 2.**
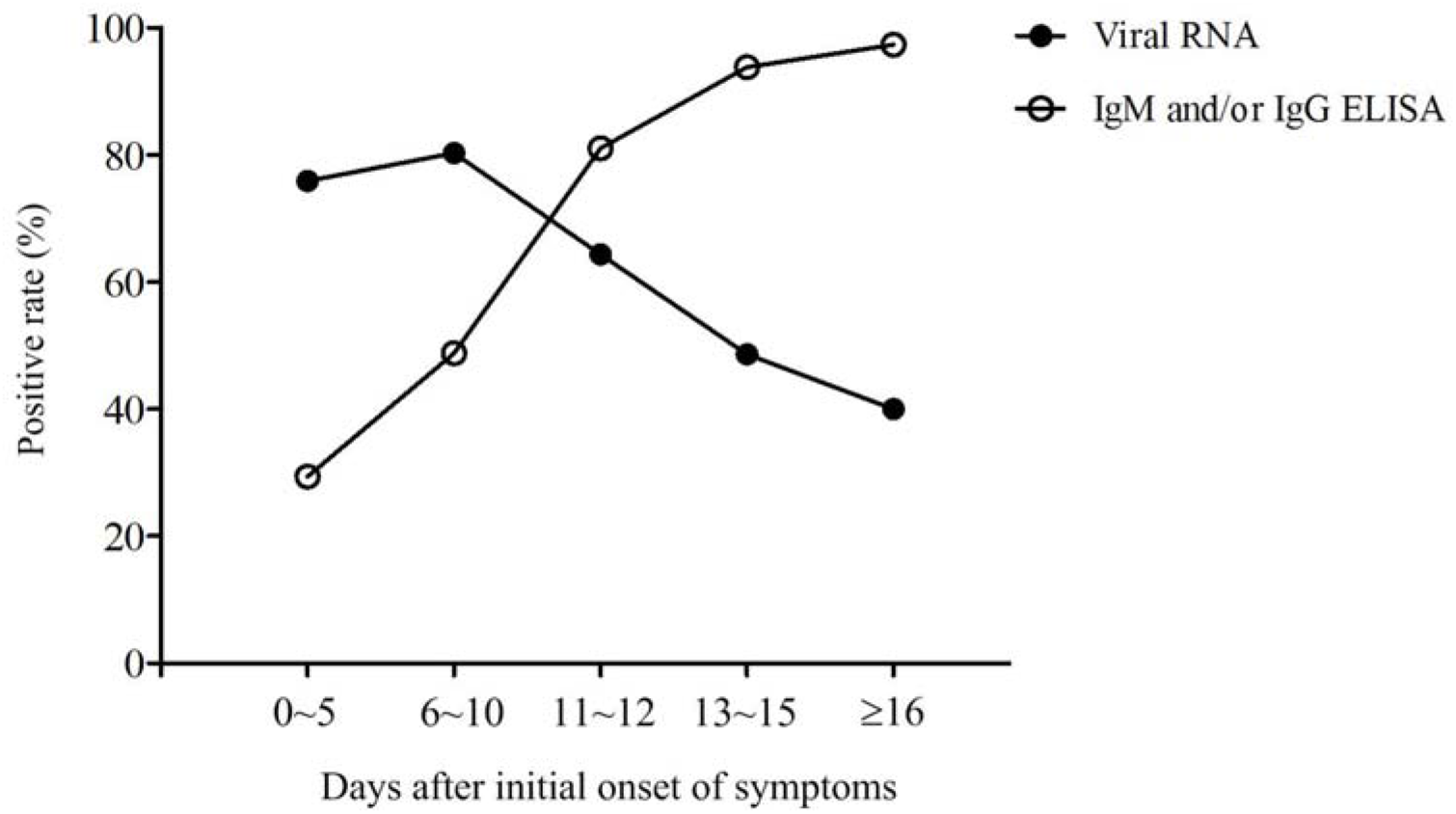
Dynamics of the positive rate of viral RNA and antibody of the patients at the different stages of disease. The disease courses were divided into five phases of 0-5, 6-10,11-12, 13-15 days and more than 16 days after initial onset of symptoms. The positive rate of viral RNA (solid circle) and antibody (hollow circle) of the patients at the different phase of disease was shown.

**Table 2.** Viral RNA and antibody positive rates of the patients detected each day from initial onset of symptoms

| Day | Viral RNA<sup>+</sup> | IgM<sup>+</sup>and/or IgG<sup>+</sup> | IgM<sup>+</sup> | IgG<sup>+</sup> | IgM<sup>+</sup>and IgG<sup>+</sup> |
| --- | --- | --- | --- | --- | --- |
| 0 | 100.0 (2/2) | 50.0 (1/2) | 0.0 (0/2) | 50.0 (1/2) | 0.0 (0/2) |
| 1 | 83.3 (5/6) | 33.3 (1/3) | 0.0 (0/3) | 33.3 (1/3) | 0.0 (0/3) |
| 2 | 71.4 (5/7) | 0.0 (0/3) | 0.0 (0/3) | 0.0 (0/3) | 0.0 (0/3) |
| 3 | 61.5 (8/13) | 25.0 (1/4) | 25.0 (1/4) | 0.0 (0/4) | 0.0 (0/4) |
| 4 | 69.2 (9/13) | 50.0 (2/4) | 0.0 (0/4) | 0.0 (0/4) | 50.0 (2/4) |
| 5 | 92.3 (12/13) | 0.0 (0/1) | 0.0 (0/1) | 0.0 (0/1) | 0.0 (0/1) |
| 6 | 62.5 (5/8) | 60.0 (3/5) | 0.0 (0/5) | 0.0 (0/5) | 60.0 (3/5) |
| 7 | 88.9 (8/9) | 50.0 (2/4) | 25.0 (1/4) | 0.0 (0/4) | 25.0 (1/4) |
| 8 | 81.8 (18/22) | 54.5 (6/11) | 18.2 (2/11) | 0.0 (0/11) | 36.4 (4/11) |
| 9 | 85.0 (17/20) | 42.9 (6/14) | 7.1 (1/14) | 7.1 (1/14) | 28.6 (4/14) |
| 10 | 75.0 (9/12) | 42.9 (3/7) | 14.3 (1/7) | 0.0 (0/7) | 28.6 (2/7) |
| 11 | 72.2 (13/18) | 81.8 (9/11) | 18.2 (2/11) | 27.3 (3/11) | 36.4 (4/11) |
| 12 | 50.0 (5/10) | 80.0 (8/10) | 30.0 (3/10) | 20.0 (2/10) | 30.0 (3/10) |
| 13 | 44.4 (4/9) | 90.9 (10/11) | 27.3 (3/11) | 9.1 (1/11) | 54.5 (6/11) |
| 14 | 69.2 (9/13) | 100.0 (17/17) | 29.4 (5/17) | 0.0 (0/17) | 70.6 (12/17) |
| 15 | 30.8 (4/13) | 90.0 (18/20) | 10.0 (2/20) | 15.0 (3/20) | 65.0 (13/20) |
| 16 | 50.0 (4/8) | 100.0 (16/16) | 12.5 (2/16) | 25.0 (4/16) | 62.5 (10/16) |
| 17 | 57.1 (4/7) | 88.9 (16/18) | 0.0 (0/18) | 5.6 (1/18) | 83.3 (15/18) |
| 18 | 33.3 (2/6) | 100.0 (16/16) | 0.0 (0/16) | 6.3 (1/16) | 93.8 (15/16) |
| 19 | 33.3 (3/9) | 100.0 (19/19) | 10.5 (2/19) | 21.1 (4/19) | 68.4 (13/19) |
| 20 | 0.0 (0/1) | 83.3 (5/6) | 0.0 (0/6) | 33.3 (2/6) | 50.0 (3/6) |
| >20 | 36.8 (7/19) | 97.2 (35/36) | 2.8 (1/36) | 8.3 (3/36) | 86.1 (31/36) |

Data are % (n/N). Day= the day after initial onset of symptoms. Viral RNA<sup>+</sup>=a positive result detected by real time RT-PCR. IgM<sup>+</sup>=a positive result detected by IgM ELISA and simultaneously a negative result detected by IgG ELISA. IgG<sup>+</sup>=a positive result detected by IgG ELISA and simultaneously a negative result detected by IgM ELISA. IgM<sup>+</sup> and IgG<sup>+</sup>=a dual positive result detected by IgM and IgG ELISA. IgM<sup>+</sup>and/or IgG<sup>+</sup>=at least a positive result detected by IgM and IgG ELISA.

**Table 3.** Viral RNA and antibody positive rates of the patients detected in different stages of disease

| Days | Viral RNA<sup>+</sup> | IgM<sup>+</sup>and/or IgG<sup>+</sup> | IgM<sup>+</sup> | IgG<sup>+</sup> | IgM<sup>+</sup>and IgG<sup>+</sup> |
| --- | --- | --- | --- | --- | --- |
| 0-5 | 75.9 (41/54) | 29.4 (5/17) | 5.9 (1/17) | 11.8 (2/17) | 11.8 (2/17) |
| 6-10 | 80.3 (57/71) | 48.8 (20/41) | 12.2 (5/41) | 2.4 (1/41) | 34.1 (14/41) |
| 11-12 | 64.3 (18/28) | 81.0 (17/21) | 23.8 (5/21) | 23.8 (5/21) | 33.3 (7/21) |
| 13-15 | 48.6 (17/35) | 93.8 (45/48) | 20.8 (10/48) | 8.3 (4/48) | 64.6 (31/48) |
| ≥16 | 40.0 (20/50) | 96.4 (107/111) | 4.5 (5/111) | 13.5 (15/111) | 78.4 (87/111) |
| Total | 64.3 (153/238) | 81.5 (194/238) | 10.9 (26/238) | 11.3 (27/238) | 59.2 (141/238) |

Data are % (n/N). Days= the day after initial onset of symptoms. Viral RNA<sup>+</sup>=a positive result detected by real time RT-PCR.
IgM<sup>+</sup>=a positive result detected by IgM ELISA and simultaneously a negative result detected by IgG ELISA. IgG<sup>+</sup>=a positive result detected by IgG ELISA and simultaneously a negative result detected by IgM ELISA. IgM<sup>+</sup> and IgG<sup>+</sup>=a dual positive result detected by IgM and IgG ELISA. IgM<sup>+</sup>and/or IgG<sup>+</sup>=at least a positive result detected by IgM and IgG ELISA.

**Table 4.** Comparison of the antibody positive rates between the confirmed and suspected patients

|  | 0-5 days | 6-10 days | ≥11 days |
| --- | --- | --- | --- |
| Confirmed | 55.6 (5/9) | 44.0 (11/25) | 93.3 (111/119) |
| Suspected | 0.0 (0/8) | 56.3 (9/16) | 95.1 (58/61) |

Data are % (n/N). Confirmed=confirmed patients. Suspected=suspected patients.

## Discussion

The outbreak of the recently emerged novel coronavirus (SARS-CoV-2) poses a challenge for public health laboratories, especially for clinical laboratories of the hospitals in Wuhan, China. Although serological assay is a frequently used method for viral infection screening and diagnosis, there are few reports about serological assay in detection of SARS-CoV-2 up to now. In this study, we report the application of the SARS-CoV-2 N protein-based ELISA for detection of IgM and IgG antibodies in the admitted hospital patients with confirmed or suspected SARS-CoV-2 infection. The results showed that the positive rates of IgM and IgG were significantly higher than that of viral RNA detected by real-time RT-PCR on the pharyngeal swab specimens of all enrolled patients. This result is further supported by the fact that the suspected patients had the same positive rate of antibody as the confirmed patients. These data strongly demonstrated that the clinically suspected patients were mostly infected by SARS-CoV-2.

In this study, the serum samples were collected in a time period of 9 days from the patients in different stages of disease. The positive rates of nucleic acid test and serological assay in total populations cannot reflect their diagnostic value in surveillance and control of the disease, because the production of antiviral antibodies will decrease the positive rates of the nucleic acid test as the disease progresses. In order to objectively determine the disease stage of the patients, we used the initial onset of symptoms of the patients as the start time point. Based on the day when the test sample was collected, all patients were defined to the different stages of disease. The resulted dynamics patterns of positive rates of viral RNA and antiviral antibodies proved the rationality of the disease stage definition. Our analysis identified the 11th day after initial onset of symptoms as a key time point in the disease process when most infected patients produce antiviral antibodies. After this time point, the diagnosis for viral infection should majorly depend on serological assay. Before this time point, nucleic acid test is important for confirmation of viral infection. The combination of serological assay can greatly improve the diagnostic efficacy.

According to the rapid advice guideline for the diagnosis and treatment of SARS-CoV-2 currently implemented in China, ^20^ a confirmed case of COVID-19 patients exclusively depends on the positive result of nucleic acid test or virus gene sequencing. Although this is a preliminary ELISA assay for SARS-CoV-2, our study strongly demonstrate that serological assay is very important for surveillance and control of the current COVID-19, especially in Wuhan of China, where a lot of patients are waiting to be confirmed at present. Moreover, most of patients had symptoms for more than 11 days.

## Data Availability

The data used to support the findings of this study are included within the article.

## Acknowledgements

This work was supported by the National Natural Science Foundation of China (81801984, 81830003); the National Key Research and Development Program of China (2019YFC130030); and the China Postdoctoral Science Foundation (2019M664008). We thank all heathy-care workers involved in this study. We thank Wuhan Institute of Virology of Chinese Academy of Sciences and Zhuhai Lizhu Diagnostics Inc. for providing assistance in ELISA detection.

## Authors’ contributions

S.Z., S.W., and L.L. conceived the study and designed experimental procedures; W.L., Y.Z., W.N.,Y.D., W.W., S.T., X.J., J.D., Q.H., Z.H., W.X., Y.Z., B.Z., Z.T., X.Z., H.L., Z.R., H.J., and X.R. collected patients’ samples. Q.W. and L.T. performed viral RNA tests. G.K.,W.L. W.N., and Y.Z established ELISA and performed serological assays. S.W., L.L., S.Z., and W.L. wrote the paper. All authors contributed to data acquisition, data analysis, or data interpretation, and reviewed and approved the final version.

